# Diagnostic Value in Discarded Reads: Re-evaluating Multi-Mapping cfDNA Fragmentomics and GC Bias in Early-Stage Cancer

**DOI:** 10.64898/2026.11.05.26367150

**Authors:** Kalyan Ram Uppaluri, Hima Jyothi Challa, Krishna Kumar Vempati, Lakshmi Nirmala Kadali

**Affiliations:** Uppaluri K&H Personalized Medicine Clinic Private Limited. (GenepoweRx), Hyderabad, India

**Keywords:** cell-free DNA, liquid biopsy, multi-mapping reads, mapping quality, satellite DNA, heterochromatin, fragmentomics, early cancer detection, matched case–control

## Abstract

**Background:** Low-coverage plasma cell-free DNA (cfDNA) sequencing is an inexpensive substrate for cancer detection. Standard pipelines discard fragments with low mapping quality, which originate mostly from repetitive DNA, although pericentromeric satellite repeats are frequently hypomethylated in cancer. We asked whether the fraction of multi-mapping cfDNA fragments differs between early-stage cancer and matched healthy controls, and how it compares with conventional fragmentomic features under strict control of technical factors.

**Methods:** We analysed public plasma cfDNA whole-genome sequencing from 25 patients with stage I–III breast cancer and 25 healthy controls matched 1:1 for age, sex, smoking and alcohol use, each pair processed in the same library and sequencing batch, and from 8 early-stage hepatocellular carcinoma (HCC) cases with 8 matched controls. For every sample we computed the fraction of autosomal fragments with MAPQ < 30 and the share of these fragments located in centromeric and heterochromatic cytobands, and related them to stage, grade and copy-number deviation as proxies of tumour burden. Classifiers were evaluated by leave-one-pair-out cross-validation in breast cancer and, trained on breast cancer only, applied unchanged to HCC. Robustness to GC content and sequencing depth was tested by within-fold residualisation and by fragment-level GC weighting. Conventional fragmentomic features (fragment length, end motifs, copy number, DELFI-like profiles and window features at transcription start sites) were evaluated under four GC-handling analyses.

**Results:** The multi-mapping fraction was higher in breast cancer than in matched controls (median 5.38 % vs 5.19 %; higher in 19/25 pairs; paired p = 0.0006; cross-validated AUC 0.81, 95 % CI 0.67–0.94), in stage I (9/10 pairs), stage II (7/10) and stage III (3/5) disease and in every sequencing batch. The excess was concentrated in centromeric and heterochromatic bands, whose share of multi-mapping fragments rose from 22.0 % to 23.4 % (19/25 pairs; p = 0.004). Both signals persisted after within-fold adjustment for GC content and/or depth (AUC 0.64–0.76 for the multi-mapping fraction, 0.71–0.76 for the heterochromatic share) and under fragment-level GC weighting (18/25 pairs; p = 0.003). The multi-mapping difference was largest in stage I and decreased with stage (Spearman ρ = −0.42, p = 0.04), and neither feature was associated with grade or copy-number deviation. In public tissue methylomes, satellites were hypomethylated in breast tumours to a similar extent as other late-replicating DNA, but the bin-level association between tumour hypomethylation and the cfDNA excess was weak and was explained by GC content. In HCC, the pre-specified endpoint, the breast-trained multi-mapping model, was not met (AUC 0.73; 5/8 pairs; sign test p = 0.36). In an exploratory analysis that was not pre-specified, the heterochromatic share was higher in 7/8 HCC pairs (p = 0.016; locked AUC 0.86, 95 % CI 0.62–1.00); across all 33 pairs, including the discovery pairs, it was higher in the cancer case in 26 (post hoc sign test p = 0.0007). In contrast, conventional fragmentomic features reached AUC 0.71 (breast) and 0.77 (HCC) only without fragment-level GC control; they tracked the per-sample coverage–GC dependence and fell to chance under each of three GC-controlled analyses, including one that preserves promoter-specific signal.

**Conclusions:** Multi-mapping cfDNA fragments, which are routinely discarded, differed between early-stage breast cancer and batch-matched controls, largely independently of GC content and sequencing depth, with an excess in centromeric and heterochromatic DNA. The signal did not scale with stage, its biological origin is unproven, and a read-quality contribution cannot be excluded from fragment files. It is a candidate marker that requires confirmation in larger cohorts with raw reads and tumour-fraction estimates.

## 1. Introduction

Cell-free DNA in plasma carries information about its cells of origin through its sequence, fragmentation pattern and genomic distribution [1–3]. In cancer, tumour-derived cfDNA differs in fragment length [4,5], fragment-end sequence [6,14] and genome-wide coverage [7], and these “fragmentomic” signals can be read from low-coverage whole-genome sequencing (WGS) without knowledge of tumour mutations [8,9]. Most analyses, however, use only uniquely mapped fragments. Fragments with low mapping quality, which derive largely from satellite and other repetitive DNA, are filtered out before any feature is computed.

There are biological reasons to look at these fragments. Pericentromeric satellite repeats are among the most consistently hypomethylated sequences in cancer, and their hypomethylation has been linked to chromatin decondensation and altered transcription [15]. Genome-wide analyses of repeat elements in cfDNA have recently shown that repeat landscapes distinguish cancer patients from healthy individuals [13]. If repeat-derived DNA is released or protected differently in cancer, the multi-mapping fraction of a cfDNA library, a quantity available from every alignment at no extra cost, could itself carry signal.

Fragmentomic measurements are also sensitive to technical factors, including blood processing, library preparation, sequencing batch and alignment [12], and to the GC content of the sequenced fragments [19]. A credible marker must therefore be evaluated in a design that removes batch differences and must be shown to be robust to GC content and sequencing depth. Here we study the multi-mapping fraction in public cohorts in which every early-stage cancer case is individually matched to a control processed in the same library and sequencing batch [11]. We test its robustness to GC content and depth, apply the breast-trained model unchanged to early-stage liver cancer, and compare it with conventional fragmentomic features evaluated under the same conditions.

## 2. Methods

### 2.1 Cohorts

#### Breast cancer cohort

Plasma cfDNA WGS fragment files for 25 patients with breast cancer and 25 matched healthy controls were obtained from the public CRAG data release (Zenodo record 6914806; CC-BY 4.0) [11]. Cases came from the University of Cincinnati Biorepository and controls from the UC Fernald Community Cohort, matched 1:1 for age, sex, smoking history and alcohol use (IRB 2012-3923 and 2012-3745) [11]. Stages were I (n = 10), II (n = 10) and III (n = 5); receptor status was ER+/PR+/HER2− in 17, triple-negative in 4 and HER2+ in 3 (Table 1).

**Table 1.** Breast cancer cohort (25 matched pairs).

| Characteristic | Value |
|---|---|
| **Stage I / II / III** | 10 / 10 / 5 |
| **ER+/PR+/HER2− · triple-negative · HER2+ · other** | 17 · 4 · 3 · 1 |
| **Age, median (range), years** | 54 (30–74) |
| **Library/sequencing batches (pairs per batch)** | 5 (4–7) |
| **Raw fragments per sample, median (range), cases** | 25.2 M (1.6–62.7) |
| **Raw fragments per sample, median (range), controls** | 30.1 M (2.1–68.9) |

According to the original study, cfDNA was extracted with the MagMAX Cell-Free DNA Isolation Kit, libraries were built from 1 ng cfDNA with KAPA HyperPrep and sequenced on an Illumina NovaSeq 6000 (PE150), and each case and its matched control were prepared and sequenced in the same batch (five batches). Reads were trimmed (Trimmomatic), aligned to GRCh37 with BWA-MEM and de-duplicated (samblaster) [11]. Labels, pairing, stage and batch were taken from CRAG Additional file 1, Supplementary Table 2.

#### Liver cancer cohort

From the same release, 8 patients with early-stage HCC (stage 1a–2) and 8 controls matched by the same criteria (5 male and 3 female pairs) were processed identically.

### 2.2 Multi-mapping fragment features

For each sample, all autosomal fragments of 50–500 bp were used at full depth (no downsampling). The **multi-mapping fraction** is the fraction of these fragments with MAPQ < 30; the fraction with MAPQ = 0 was also recorded. Each fragment midpoint was assigned to a UCSC hg19 cytoband, and bands with stain acen, gvar or stalk were defined as centromeric/heterochromatic. The **heterochromatic share** is the fraction of MAPQ < 30 fragments whose midpoints fall in these bands. We also computed the fractions of all fragments that are multi-mapping inside and outside these bands, the share of uniquely mapped (MAPQ ≥ 30) fragments in these bands, and the median length of multi-mapping fragments. Because BWA-MEM assigns MAPQ 0 to reads with equally good alternative alignments and intermediate values (1–29) when alternatives are only slightly worse or the alignment is uncertain, the MAPQ < 30 fraction was also split into MAPQ 0 and MAPQ 1–29 components.

### 2.3 Technical covariates

Sequencing depth was the raw number of fragments per sample, counted before any filter. The per-sample dependence of coverage on GC content (GC slope) was computed as the slope of median-normalised 1-Mb coverage of MAPQ ≥ 30 fragments against bin GC content (bins with < 5 % N; central 98 % of coverage). For fragment-level GC weighting [19], each fragment received a weight that equalises the sample’s joint distribution of fragment length (10-bp bins) and fragment GC content (1 % bins) to a single genome-derived expected distribution, identical for all samples and independent of labels (weights clipped to [0.2, 5], set to 1 in bins with < 20 fragments, rescaled to mean 1); the multi-mapping fraction was then recomputed as a weighted ratio.

### 2.4 Conventional fragmentomic features

For comparison, the following features were computed from MAPQ ≥ 30 autosomal fragments downsampled to 1.4 million per sample (the depth of the shallowest sample): fragment length (6 features), 5′ end motif (motif diversity score, CCCA frequency, fraction of C-starting ends) [6], arm-level copy-number deviation, DELFI-like short/long profile deviation [7], and seven window features at 10,286 transcription start sites (TSSs) following Wang et al. [10] (IFS, EDI, FDI, WPS, OCF [18], PFE, mtDNA ratio). Copy-number and DELFI features used bin-level GC correction and healthy references rebuilt inside each cross-validation fold. These features were evaluated under four GC-handling analyses: (i) bin-level GC correction only; (ii) fragment-level GC weighting of every feature (Section 2.3); (iii) GC-internal relative features at 1.4 M fragments, in which each feature is expressed relative to a GC-matched reference inside the same sample (C-end fraction minus the C fraction at interior positions +5 to +14 of the same fragments; CCCA log-enrichment and end-motif Kullback–Leibler divergence relative to interior 4-mers; IFS and FDI at TSSs divided by the same features at GC-matched non-TSS windows); and (iv) the same GC-internal features at 10 M fragments in the 19 breast pairs and 8 liver pairs with sufficient depth. Analyses (iii) and (iv) followed a protocol written before they were run.

### 2.5 Tumour-burden proxies

The public metadata contain stage, grade and receptor status but no tumour size, ctDNA fraction or somatic variant data, and tumour-fraction estimation from low-coverage data of early-stage disease is unreliable. We therefore used three proxies: stage (I–III, ordinal), grade (G1–G3) and the copy-number deviation score (root-mean-square arm-level z-score against controls, Section 2.4). Within breast cancer pairs, the case–control difference of each multi-mapping feature was correlated with stage and grade (Spearman), and case values were correlated with the copy-number score.

### 2.6 Comparison with public tissue methylation maps

To check whether the compartments carrying the cfDNA excess are hypomethylated in tumour tissue, we used public hg19 whole-genome bisulfite sequencing (WGBS) data: five TCGA breast tumours and one adjacent normal breast sample (reprocessed by Zhou et al. [20]), and Roadmap Epigenomics adult liver (E066 [21]). No public HCC WGBS was available in these resources, so liver is a normal-tissue baseline only. The autosomes were divided into six mutually exclusive compartments, assigned in this order: alpha satellite (RepeatMasker ALR/Alpha [22]), other satellites (all other RepeatMasker Satellite elements), the remaining centromeric and heterochromatic cytobands, common partially methylated domains (PMDs) and common highly methylated domains (HMDs) [20], and the rest of the genome. Common PMDs, which lose methylation in most cancers [20], served as a positive control. For each tissue sample we computed mean CpG methylation per compartment (CpGs with depth ≥ 4). For each cfDNA sample we computed, per compartment, the low-MAPQ fraction and the compartment’s share of all low-MAPQ fragments, and compared cases with matched controls. The single pre-specified primary test was a Spearman correlation across 1-Mb autosomal bins between the mean breast tumour − normal methylation difference and the median paired case − control difference in the low-MAPQ fraction; a negative correlation was predicted. The protocol was written before the external files were opened. Because both quantities depend on GC content, a GC-adjusted partial correlation and a within-chromosome circular-shift permutation (preserving spatial autocorrelation) were added post hoc.

### 2.7 Evaluation

All comparisons are within matched pairs. Paired differences were tested with Wilcoxon signed-rank tests and one-sided exact sign tests. Classifiers were L2-regularised logistic regression (scikit-learn [17]; C = 1; features standardised within each training fold). In breast cancer, performance was estimated by **leave-one-pair-out (LOPO)** cross-validation, holding out each case with its matched control; we report AUC with bootstrap 95 % CI (2,000 resamples), sensitivity at 95 % specificity, and sensitivity and specificity at the classifier’s default decision threshold. For robustness, the multi-mapping features were residualised within each training fold on the GC slope, on depth, or on both, and the model was refitted. Conventional fragmentomic models were additionally assessed with 1,000 (200 for analysis ii) within-pair label permutations. For the liver cohort, models were trained once on all 50 breast samples, locked, and applied unchanged; endpoints were AUC with a pair-level bootstrap 95 % CI and the number of pairs in which the case scored above its control.

#### Pre-specification

The multi-mapping fraction was first identified in exploratory analysis of the breast cohort, so the breast results are discovery results. Before the liver data were unpacked, a protocol fixed the locked multi-mapping model (M2) and the 11-feature fragmentomic model (M1) as liver endpoints. The heterochromatic share was not pre-specified for liver; its liver analysis is exploratory and hypothesis-generating, and is not a replication. The pooled sign tests across both cohorts were defined post hoc and include the discovery pairs, so they describe consistency of direction rather than independent confirmation.

## 3. Results

### 3.1 A batch-matched cohort

All 50 breast-cohort samples passed processing (1.6–68.9 million raw fragments; Table 1). Raw fragments are all records in each public fragment file, before any filter on chromosome, length or MAPQ (Table S6). Raw fragment counts did not differ between cases and controls (median 25.2 vs 30.1 M; higher in the case in 12/25 pairs; paired p = 0.30), and neither did those in the liver cohort (median 26.7 vs 28.8 M; 4/8 pairs; p = 1.0). Every comparison is between a case and a control from the same library and sequencing batch. These counts measure sequencing depth, which depends on library input and sequencing allocation, not the amount of cfDNA in plasma. Plasma cfDNA concentration was not available in the public metadata, so it could not be compared. For this reason all multi-mapping measures are expressed as fractions of each sample’s fragments, not as counts.

### 3.2 Multi-mapping fragments are more frequent in early breast cancer

The fraction of autosomal cfDNA fragments with MAPQ < 30 was higher in breast cancer than in matched controls (median 5.38 % vs 5.19 %; higher in 19/25 pairs; paired p = 0.0006; univariate AUC 0.83; Fig. 1a, Table 2). Fragments with MAPQ = 0 showed the same difference (21/25 pairs; p = 0.0008). As a single-feature classifier, the multi-mapping fraction gave a LOPO AUC of 0.81 (95 % CI 0.67–0.94), with sensitivity 0.68 and specificity 0.84; sensitivity at 95 % specificity was 0.12.

**Figure 1.**
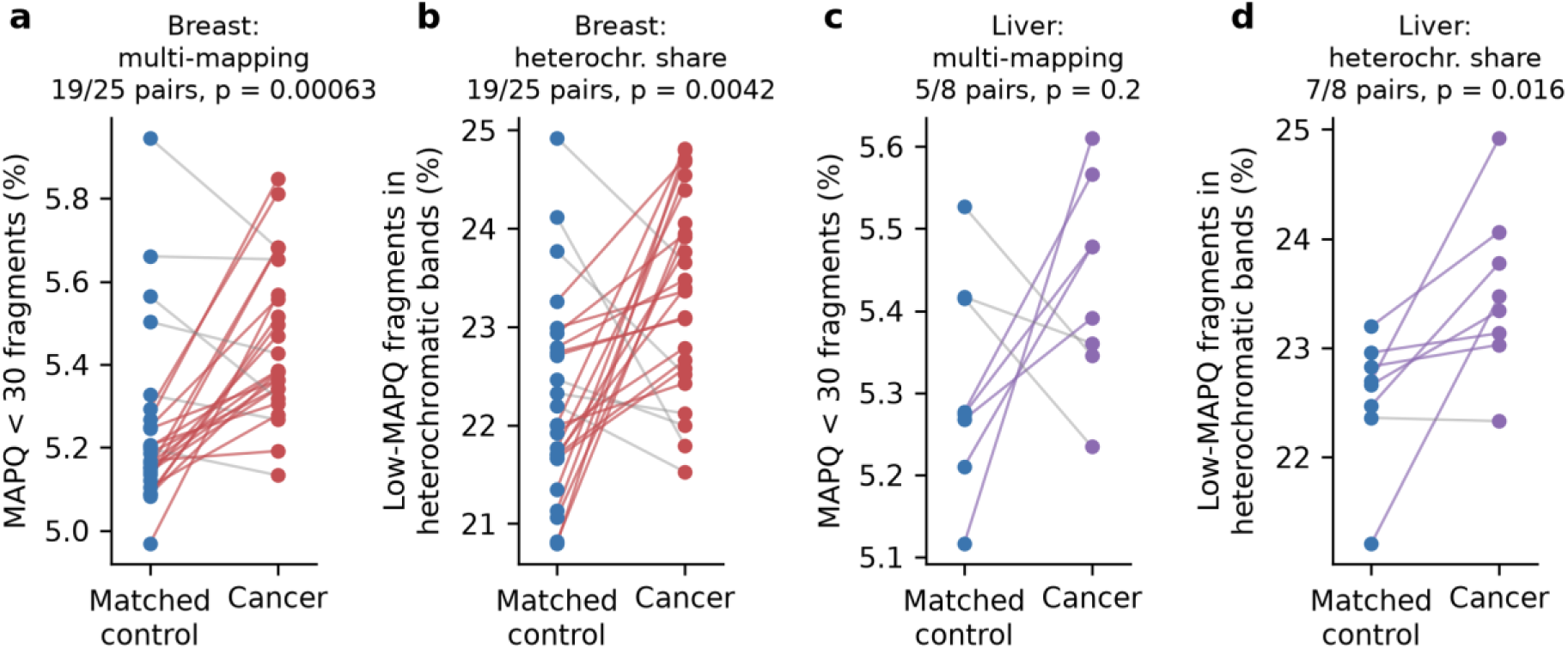
Multi-mapping cfDNA fragments in matched pairs. (a, c) Percentage of autosomal fragments with MAPQ < 30 in each cancer case and its matched control, breast (a) and liver (c). (b, d) Percentage of MAPQ < 30 fragments located in centromeric/heterochromatic cytobands (acen, gvar, stalk). Coloured lines, case > control; grey, case ≤ control; p, paired Wilcoxon test.

**Table 2.** Multi-mapping fragment features in matched pairs.

| Feature | Breast: case vs control (median) | Breast pairs case > control | Breast p | Liver: case vs control (median) | Liver pairs case > control | Liver p |
|---|---|---|---|---|---|---|
| **Multi-mapping fraction (MAPQ < 30), %** | 5.38 vs 5.19 | 19/25 | 0.0006 | 5.43 vs 5.28 | 5/8 | 0.20 |
| **MAPQ = 0 fraction, %** | 3.28 vs 3.16 | 21/25 | 0.0008 | 3.31 vs 3.21 | 5/8 | 0.11 |
| **Multi-mapping, in heterochromatic bands, % of all** | 1.28 vs 1.15 | 20/25 | 0.0004 | 1.28 vs 1.20 | 5/8 | 0.11 |
| **Multi-mapping, outside heterochromatic bands, % of all** | 4.17 vs 4.04 | 18/25 | 0.002 | 4.14 vs 4.12 | 4/8 | 0.46 |
| **Heterochromatic share of multi-mapping fragments, %** | 23.4 vs 22.0 | 19/25 | 0.004 | 23.4 vs 22.7 | **7/8** | **0.016** |
| **Median length of multi-mapping fragments, bp** | 166 vs 165 | 14/25 | 0.42 | 165 vs 166 | 3/8 | 0.81 |

p, paired Wilcoxon signed-rank test (two-sided). Liver rows other than the multi-mapping fraction are exploratory (not pre-specified).

The difference was present at every stage, with the case higher than its control in 9/10 stage I, 7/10 stage II and 3/5 stage III pairs, and in every library batch (5/6, 4/7, 3/4, 4/4 and 3/4 pairs). The median length of multi-mapping fragments did not differ between groups (166 vs 165 bp; p = 0.42), so the effect is not a by-product of shorter fragments. The excess was shared proportionally between the two mapping-quality classes: MAPQ 0 (3.28 % vs 3.16 %; 21/25 pairs; p = 0.0008) and MAPQ 1–29 (2.14 % vs 2.04 %; 19/25 pairs; p = 0.0006), and the proportion of MAPQ 0 among multi-mapping fragments did not differ (60.7 % vs 61.0 %; 10/25 pairs; p = 0.08).

### 3.3 The excess localises to centromeric and heterochromatic DNA

The multi-mapping excess was strongest in centromeric and heterochromatic bands. The fraction of all fragments that were multi-mapping and located in these bands was higher in 20/25 pairs (p = 0.0004), compared with 18/25 pairs (p = 0.002) outside them. As a result, the share of multi-mapping fragments located in centromeric and heterochromatic bands rose from 22.0 % in controls to 23.4 % in cases (19/25 pairs; p = 0.004; LOPO AUC 0.76, 95 % CI 0.62–0.89; Fig. 1b). Uniquely mapped fragments also shifted towards these bands (15/25 pairs; p = 0.026).

### 3.4 Robustness to GC content and sequencing depth

Across breast samples, the multi-mapping fraction was correlated with sequencing depth (Spearman ρ = −0.68) and with the GC slope (ρ = 0.49), and the heterochromatic share less so (ρ = −0.52 and 0.30). Depth did not differ between cases and controls (above). Adjusting for these covariates within each training fold reduced but did not remove the signal (Fig. 2a). The multi-mapping fraction gave LOPO AUC 0.64 after adjustment for the GC slope, 0.76 after adjustment for depth and 0.76 after adjustment for both. The heterochromatic share was less affected (0.71, 0.76 and 0.76). With fragment-level GC weighting, the multi-mapping fraction remained higher in cancer (median 5.34 % vs 5.16 %; 18/25 pairs; p = 0.003; LOPO AUC 0.78).

**Figure 2.**
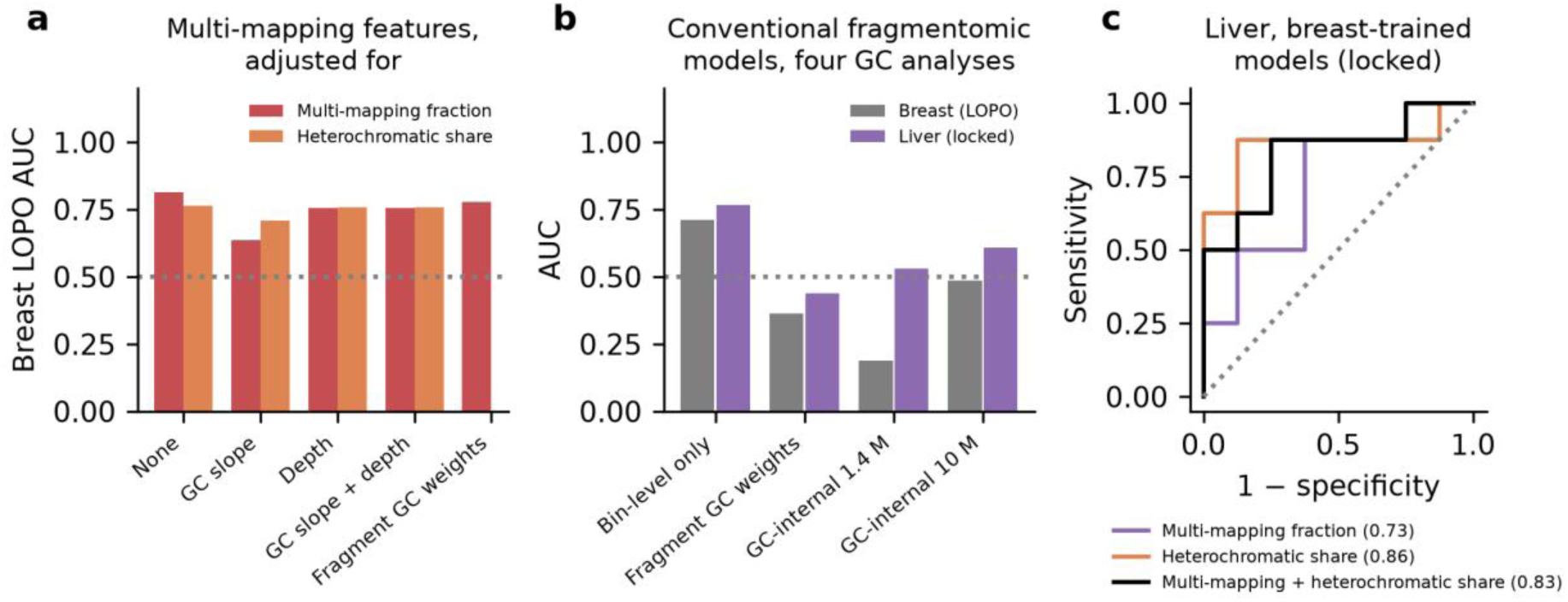
Robustness and comparison with conventional fragmentomic features. (a) Breast leave-one-pair-out AUC of the multi-mapping fraction and the heterochromatic share without adjustment, after within-fold residualisation on the GC slope, on depth or on both, and (multi-mapping fraction) with fragment-level GC weights. (b) AUC of the conventional fragmentomic model (11 features; 5 relative features in the GC-internal analyses) in breast (leave-one-pair-out) and liver (locked) under the four GC-handling analyses. (c) ROC curves of breast-trained multi-mapping models applied unchanged to liver cancer. Dotted lines, AUC 0.5.

### 3.5 Relation to tumour-burden proxies

The multi-mapping signal did not increase with disease extent. The case–control difference in the multi-mapping fraction was largest in stage I (median 0.36 percentage points), smaller in stage II (0.17) and close to zero in stage III (0.02; Spearman ρ with stage −0.42, p = 0.04). The difference in the heterochromatic share showed no trend with stage (median 1.23, 0.70 and 0.86 percentage points; ρ = −0.23, p = 0.27). Neither difference was associated with grade (ρ = −0.01 and 0.10; 24 pairs with grade recorded), and neither feature was associated with the copy-number deviation score in cases (ρ = 0.23, p = 0.27; ρ = 0.16, p = 0.44). These data therefore do not show that the signal scales with tumour burden; with 5 stage III pairs and no direct tumour-fraction measurement, they also cannot exclude it.

### 3.6 Comparison with public tissue methylation maps

Breast tumours were hypomethylated relative to the adjacent normal sample in every compartment (Table S5). The loss was similar in satellites and in the common-PMD positive control. Mean tumour − normal differences were −0.081 in alpha satellite (range across tumours −0.157 to +0.009), −0.115 in other satellites (−0.163 to −0.026), −0.101 in the remaining heterochromatic bands and −0.113 in common PMDs, compared with −0.050 in common HMDs. Satellites were well methylated in normal breast (alpha 0.58, other satellites 0.76) and normal liver (0.63 and 0.70), so they have methylation to lose. WGBS covered only about 32,000 alpha-satellite CpGs per sample, compared with about 9 million in common PMDs, because satellites map poorly with short reads.

In breast cfDNA, the increase in the heterochromatic share came from non-alpha satellites (share of low-MAPQ fragments 17.4 % vs 16.2 %; 18/25 pairs; p = 0.022), not from alpha satellite (15.3 % vs 14.7 %; 15/25; p = 0.21) or the remaining heterochromatic bands (6/25). The low-MAPQ fraction was also higher in cases inside common PMDs (3.69 % vs 3.56 %; 20/25; p = 0.002) and common HMDs (2.44 % vs 2.41 %; 21/25; p = 0.0005), so part of the excess is genome-wide. In liver cfDNA the pattern differed: the low-MAPQ fraction within alpha satellite was higher in 7/8 pairs (p = 0.035), whereas the non-alpha satellite share was lower in 5/8; the low-MAPQ fraction within common PMDs was higher in 8/8 pairs (p = 0.004).

The pre-specified bin-level test was in the predicted direction but weak: across 2,733 bins, the cfDNA excess was larger where breast tumours lost more methylation (Spearman ρ = −0.056, p = 0.004; within-chromosome circular-shift p = 0.006). The result was similar after excluding satellite- and heterochromatin-dominated bins (ρ = −0.050, p = 0.009). In the post hoc check, however, both quantities depended on bin GC content (cfDNA excess ρ = −0.39; methylation difference ρ = 0.14). After adjustment for GC, the association disappeared (partial ρ = 0.000, p = 0.99). The public maps are therefore compatible with satellite hypomethylation in breast tumours. They do not show a GC-independent link between where tumours lose methylation and where the cfDNA excess occurs.

### 3.7 Exploratory analysis in early-stage liver cancer

The pre-specified liver endpoint was not met. The multi-mapping fraction pointed in the same direction but did not differ significantly (5.43 % vs 5.28 %; 5/8 pairs; p = 0.20), and the breast-trained model M2 gave an AUC of 0.73 (95 % CI 0.36–1.00; 5/8 pairs; sign test p = 0.36), with sensitivity 0.88 and specificity 0.63 at the breast-derived threshold (Fig. 2c, Table 3).

**Table 3.** Breast-trained models applied unchanged to liver cancer (8 HCC / 8 matched controls). Only M2 was pre-specified.

| **Model** | **AUC (95 % CI)** | **Pairs case > control** | **Sign test p** |
|---|---|---|---|
| **M2 multi-mapping fraction (pre-specified)** | 0.73 (0.36–1.00) | 5/8 | 0.36 |
| **Heterochromatic share (exploratory)** | **0.86 (0.62–1.00)** | **7/8** | **0.035** |
| **Multi-mapping + heterochromatic share (exploratory)** | 0.83 (0.50–1.00) | 5/8 | 0.36 |

In an exploratory analysis that was not pre-specified, the heterochromatic share was higher in the HCC case than in its matched control in 7/8 pairs (23.4 % vs 22.7 %; p = 0.016; Fig. 1d), and a breast-trained, locked model on this feature alone gave AUC 0.86 (95 % CI 0.62–1.00; 7/8 pairs; one-sided sign test p = 0.035). The heterochromatic share was not correlated with depth (ρ = −0.04) or the GC slope (ρ = 0.19) in the liver cohort. Across all 33 matched pairs, the cancer case had the higher heterochromatic share in 26 (post hoc one-sided sign test p = 0.0007) and the higher multi-mapping fraction in 24 (p = 0.007). Because these counts include the 25 discovery pairs, they summarise consistency of direction and are not independent confirmation. With 8 pairs, the liver result generates a hypothesis for a pre-specified test in a new cohort.

### 3.8 Conventional fragmentomic features do not survive GC control

Without fragment-level GC control (analysis i), conventional fragmentomic features appeared to separate the groups. The 11-feature model gave a breast LOPO AUC of 0.71 (95 % CI 0.56–0.86; permutation p = 0.019), the end-motif family 0.76 and the window features 0.68, and the locked 11-feature model gave AUC 0.77 in HCC (8/8 pairs). The fraction of C-starting ends (18/25 pairs; p = 0.003), FDI (18/25; p = 0.004) and IFS (18/25; p = 0.014) were higher in cancer (Figs. S2, S3). These features, however, tracked the per-sample coverage–GC dependence, which was stronger in the cancer case in 21/25 breast and 8/8 liver pairs (Spearman ρ with the GC slope 0.54–0.83 for the C-end fraction, 0.78–0.79 for FDI and 0.71–0.82 for IFS; Fig. S1).

Under each of the three GC-controlled analyses, the conventional models fell to chance (Table 4; Fig. 2b). With fragment-level GC weighting, no feature differed between cases and controls (all q > 0.7), and the 11-feature model gave AUC 0.36 in breast (permutation p = 0.84) and 0.44 in HCC. The GC-internal relative features were not correlated with the GC slope (|ρ| ≤ 0.32) and carried no signal at 1.4 M fragments (breast AUC 0.19, permutation p = 0.99; HCC 0.53) or at 10 M fragments (breast 0.48, permutation p = 0.52; HCC 0.61). At both depths, the uncorrected differences reappeared to the same extent in the GC-matched references: the C fraction at interior fragment positions was higher in the case in 22/25 breast and 8/8 liver pairs, and IFS and FDI at GC-matched non-TSS windows in 18–20/25 breast and 8/8 liver pairs. AUCs below 0.5 are expected under leave-one-pair-out cross-validation when features carry no signal.

**Table 4.** Conventional fragmentomic models under four GC-handling analyses (AUC; breast LOPO / liver locked).

| Analysis | 11 features (or 5 relative) | End motif | Window |
|---|---|---|---|
| **(i) Bin-level GC correction only** | 0.71 / 0.77 | 0.76 / 0.84 | 0.68 / 0.72 |
| **(ii) Fragment-level GC weighting** | 0.36 / 0.44 | 0.36 / 0.31 | 0.34 / 0.47 |
| **(iii) GC-internal relative, 1.4 M** | 0.19 / 0.53 | 0.34 / 0.56 | 0.23 / 0.41 |
| **(iv) GC-internal relative, 10 M (19 + 8 pairs)** | 0.48 / 0.61 | 0.52 / 0.52 | 0.44 / 0.83* |
| **Multi-mapping fraction, for reference** | 0.81 / 0.73 | — | — |

*One of about 20 secondary model results across analyses (iii) and (iv); the same model gave 0.44 in breast*

## 4. Discussion

In batch-matched early-stage breast cancer, the fraction of cfDNA fragments with low mapping quality was higher in cancer than in matched controls, and the excess was concentrated in centromeric and heterochromatic DNA. The signal was present at stage I, in every sequencing batch, after adjustment for GC content and depth, and under fragment-level GC weighting. The pre-specified liver endpoint, the breast-trained multi-mapping model, was not met. In an exploratory liver analysis, the share of multi-mapping fragments falling in centromeric and heterochromatic bands was higher in 7 of 8 pairs; this is a hypothesis for a pre-specified test in an independent cohort, not a replication. Conventional fragmentomic features, by contrast, separated the groups only when fragment GC composition was left uncontrolled.

### Possible biological origin

The localisation of the excess suggests, but does not establish, a contribution from repetitive pericentromeric DNA. Pericentromeric satellites are hypomethylated in many cancers, and this has been linked to chromatin decondensation and satellite transcription [15]; genome-wide repeat profiling of cfDNA has shown that repeat landscapes differ between cancer patients and healthy individuals [13]. One hypothesis consistent with this literature is that decondensed satellite chromatin is released or protected differently during cell death, increasing the representation of satellite-derived fragments in plasma. We did not measure methylation or chromatin accessibility in these samples, and three observations argue for caution. First, the multi-mapping difference was largest in stage I and smallest in stage III, and neither feature was associated with copy-number deviation, which is the opposite of what a purely tumour-derived signal would be expected to show as tumour burden rises. The signal may instead reflect non-tumour sources, such as a systemic or immune response to early disease, or pre-analytical factors. Second, the uniquely mapped (MAPQ ≥ 30) fragments also shifted slightly towards heterochromatic bands, which is consistent with a change in genomic origin, but the effect was small (15/25 pairs). Third, public tissue methylomes gave only indirect and weak support. Satellites were hypomethylated in breast tumours, but no more than common PMDs. The breast and liver excesses involved different satellite families, and the bin-level association between tumour hypomethylation and the cfDNA excess was explained by GC content. These tissue maps come from different patients and cover few satellite CpGs, so they cannot test the hypothesis directly. A direct test requires methylation-aware cfDNA data (bisulfite or enzymatic methylation sequencing) from the same samples.

### Technical alternatives

Mapping quality depends on the genomic origin of a fragment but also on read-level properties such as base quality, sequencing errors, read length and residual adapter sequence. The open data release used here contains aligned fragment coordinates and mapping quality but not reads or base qualities, so these factors could not be measured. Two observations from the fragment files are relevant but not decisive. The excess was distributed proportionally between MAPQ 0 fragments, which typically indicate equally good alternative alignments, and MAPQ 1–29 fragments, which also arise from sequencing errors in near-repetitive sequence, so the MAPQ classes do not discriminate between the two explanations. The multi-mapping fraction is also almost the complement of the overall MAPQ ≥ 30 fraction (ρ = −0.97) and decreased with sequencing depth. The heterochromatic share is less exposed to a uniform read-quality effect, because it compares fragments within the multi-mapping set, and it was unrelated to depth and GC slope in the liver cohort. A definitive test requires re-alignment of raw reads with per-base Phred scores, mismatch rates, adapter-trimming statistics and read-length distributions compared between cases and controls.

### Does GC control remove real biology?

Fragment-level GC weighting equalises each sample’s fragment GC distribution to a genome-wide expectation. If cancer-derived cfDNA genuinely differs in GC composition, or if promoter- and CpG-island-rich regions carry a real fragmentation signal, part of that signal would be removed, so this correction alone could over-correct. For this reason we also used GC-internal features, which compare TSSs with non-promoter windows of the same GC content inside the same sample and therefore preserve promoter-specific differences. These features were also null, and the uncorrected case–control differences appeared to the same extent at GC-matched non-promoter windows and at interior fragment positions as at TSSs and fragment ends. The shift that drove the conventional features is therefore sample-wide rather than promoter- or cleavage-specific. A sample-wide GC shift could itself have a biological component, but it would not be specific to end motifs or TSS fragmentation. The multi-mapping features are not immune to GC: the multi-mapping fraction lost part of its performance after adjustment for the GC slope (LOPO AUC 0.81 to 0.64), whereas the heterochromatic share was less affected (0.76 to 0.71). We recommend that fragmentomic studies report the per-sample GC slope, test it between groups, and evaluate candidate features with both fragment-level and GC-matched internal controls.

### Sample size and analysis history

The cohorts are small (25 and 8 pairs). The multi-mapping fraction was discovered in the breast cohort, so the breast estimates are likely optimistic. The pooled 33-pair sign tests include the discovery pairs and were defined post hoc. Several GC-handling analyses of the conventional features were run, and all are reported (Table 4; Supplementary Note 2).

### Next steps

A pre-specified test of the multi-mapping fraction and the heterochromatic share in larger, independent matched cohorts across cancer types, with raw reads and base-quality metrics; repeat-resolved quantification (satellite families, alpha-satellite higher-order repeats) with repeat-aware alignment; methylation-aware sequencing of the same cfDNA, with long-read or repeat-aware methods to resolve satellite methylation; and calibration against tumour fraction from deeper sequencing or tumour-informed assays.

### Minimal residual disease

These data do not address minimal residual disease (MRD) detection. The cohort contains only pre-treatment samples, with no serial sampling, recurrence outcomes or tumour-fraction estimates. The multi-mapping signal did not increase with stage or copy-number deviation, which an MRD marker would need to do. Because MRD assays compare a patient with their own baseline, within-person serial sampling would remove the between-person variation that limits this small-effect marker. Its relevance to MRD could therefore be tested in pre- and post-treatment samples from the same patients, with depth-matched sequencing, tumour-informed ctDNA measurements as reference and follow-up for recurrence. Until then, no MRD application should be inferred.

### Limitations and proof-of-concept status

This study represents a proof-of-concept discovery rather than a ready-to-use clinical test. The cohorts are relatively small (n=25 breast pairs and n=8 liver pairs) and rely on the retrospective analysis of public data. Because the multi-mapping fraction was discovered in the breast cohort, the performance estimates in breast cancer are likely optimistic, and the pooled 33-pair sign tests were defined post hoc. Furthermore, the public data release provided aligned fragment coordinates but lacked raw sequencing reads and per-base quality scores. Consequently, we cannot completely exclude the possibility that subtle machine read errors or pre-analytical technical factors contribute to the signal. Several GC-handling analyses of the conventional features were run, and all are reported here to ensure transparency (Table 4) .Establishing the true clinical utility and biological origin of this marker will require prospective validation in larger, independent patient cohorts using raw sequence data.

## 5. Conclusion

By repurposing routinely discarded low-mapping-quality cfDNA fragments (MAPQ < 30), we demonstrate that multi-mapping reads from centromeric and heterochromatic regions are significantly elevated in early-stage breast cancer. Crucially, this robust marker survives rigorous GC-bias corrections that reduce conventional fragmentomic features (such as lengths and motifs) to random chance, exposing a major vulnerability in standard liquid biopsy pipelines.

Unconventionally, the multi-mapping excess peaks in Stage I disease and does not scale with tumor burden, suggesting it may reflect an early systemic response rather than direct tumor shedding. Extracted at zero additional cost from standard whole-genome sequencing, this GC-resistant marker challenges current filtering practices. While its exact biological origin requires confirmation and the liver cancer endpoint remains exploratory, it offers a highly accessible candidate for early cancer detection that warrants validation in larger independent cohorts.

## Supporting information

S1_breast_samples

S2_liver_Samples

covariance_adjustment

multimapping_Samples

methyl.._Crossref

raw_counts

## Data Availability

This study used only publicly available data. Plasma cfDNA fragment files are available from the CRAG data release (Zenodo record 6914806). Sample labels, pairing, stage and batch are from Additional file 1, Supplementary Table 2 of the original CRAG publication. Whole-genome bisulfite data for TCGA breast samples and common PMD/HMD coordinates are available from Zhou et al., adult liver methylation data from Roadmap Epigenomics (E066), and annotations from UCSC (hg19). All data produced in the present study are contained in the manuscript and its supplementary tables.

https://zenodo.org/records/6914806

https://zwdzwd.github.io/pmd

https://egg2.wustl.edu/roadmap/data/byDataType/dnamethylation/

https://hgdownload.soe.ucsc.edu/goldenPath/hg19/

## Declarations

### Data availability

Fragment files: Zenodo record 6914806 (CRAG; https://zenodo.org/records/6914806). Public methylation and annotation data (hg19): TCGA BRCA WGBS beds reprocessed by Zhou et al. (https://zwdzwd.github.io/pmd; samples A04X, A07I, A0CE, A0YG, A15H, NA0CE), common PMD/HMD coordinates from the same source, Roadmap Epigenomics E066 fractional methylation (https://egg2.wustl.edu/roadmap/data/byDataType/dnamethylation/), and UCSC hg19 RepeatMasker. Sample labels, pairing, stage and batch: CRAG Additional file 1, Supplementary Table 2 [11]. Per-sample multi-mapping values are in Table S4, the methylation cross-reference in Table S5, raw fragment counts in Table S6 and other features in Tables S1–S3.

### Code availability

The analysis code and study protocols will be made publicly available in a GitHub repository during peer review. https://github.com/Nirmala-k/Multi-mapping-cell-free-DNA-fragments-

### Ethics

Secondary analysis of publicly available, de-identified data. Sample collection in the source study was approved by the University of Cincinnati IRB (2012-3923, 2012-3745) with written informed consent [11].

## Acknowledgements

We thank Zhou, Zheng, Fu, Liu and colleagues for making the CRAG matched cohorts publicly available.

## Use of AI tools

Manuscript drafting were assisted by an AI model (Claude, Anthropic).

## Figure legends

**Figure S1.**
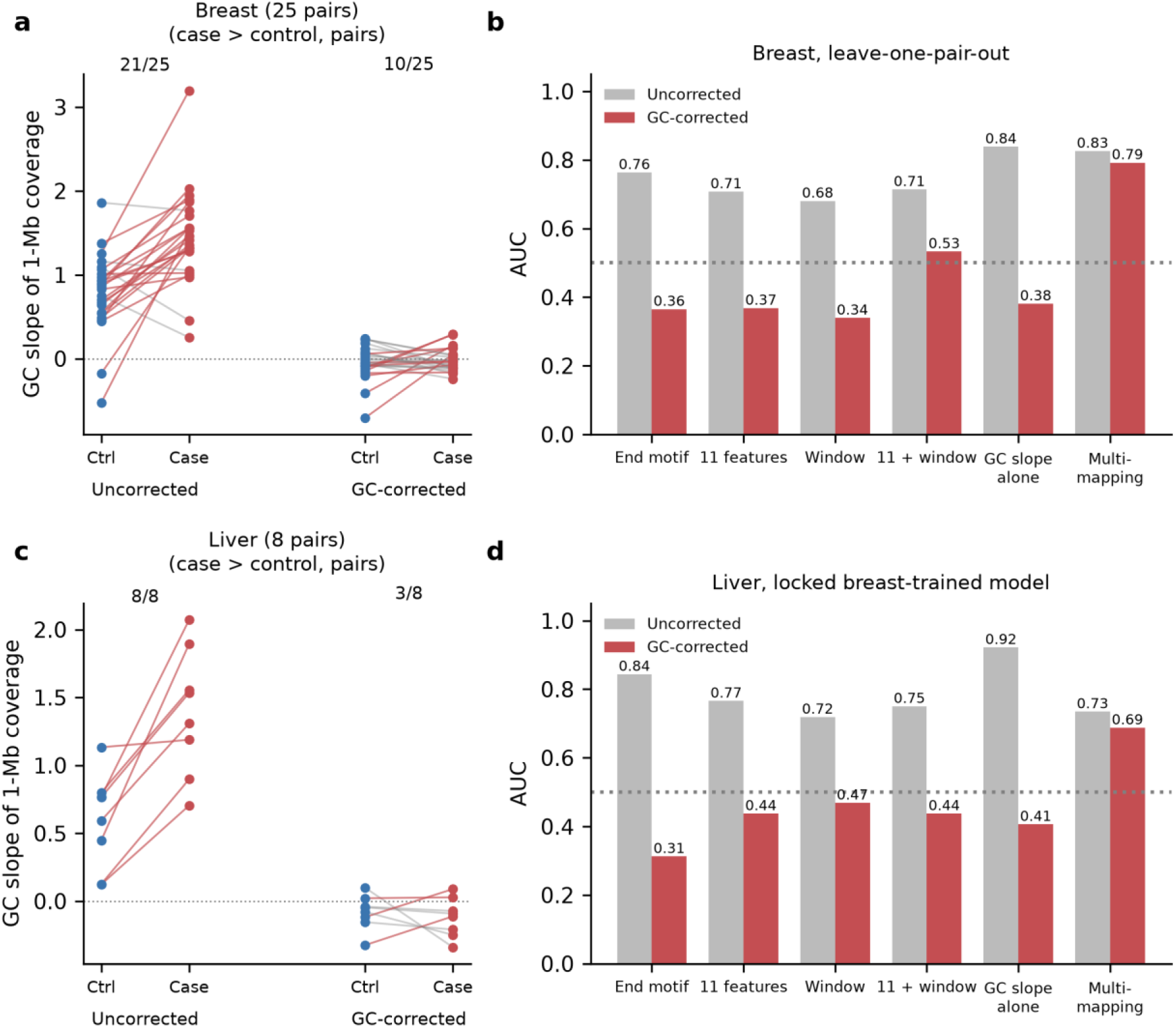
Conventional fragmentomic models with and without fragment-level GC weighting. (a, b) GC slope of 1-Mb coverage for each case and its matched control, without and with weighting, in breast (a) and liver (b). (c, d) AUC of each feature set without (grey) and with (red) weighting in breast (c) and liver (d).

**Figure S2.**
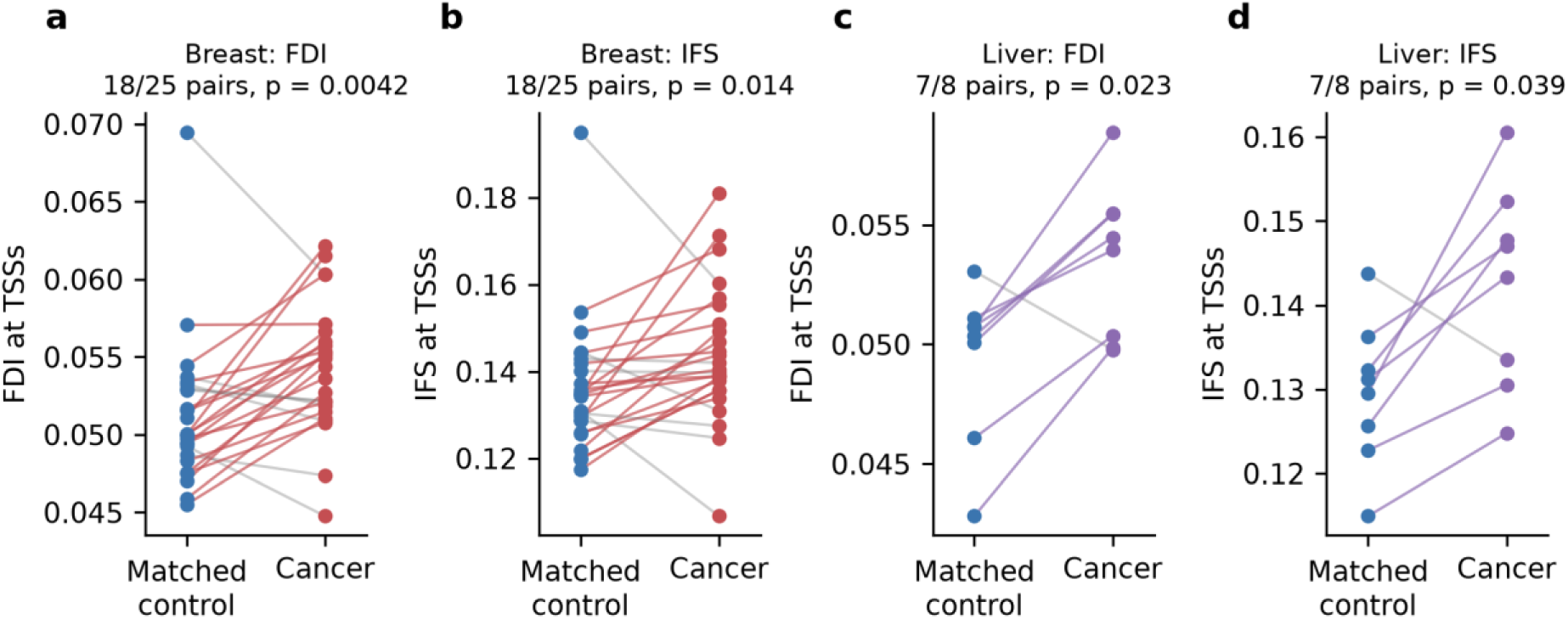
Window features at TSSs without fragment-level GC control. FDI and IFS for each case and matched control, breast (a, b) and liver (c, d).

**Figure S3.**
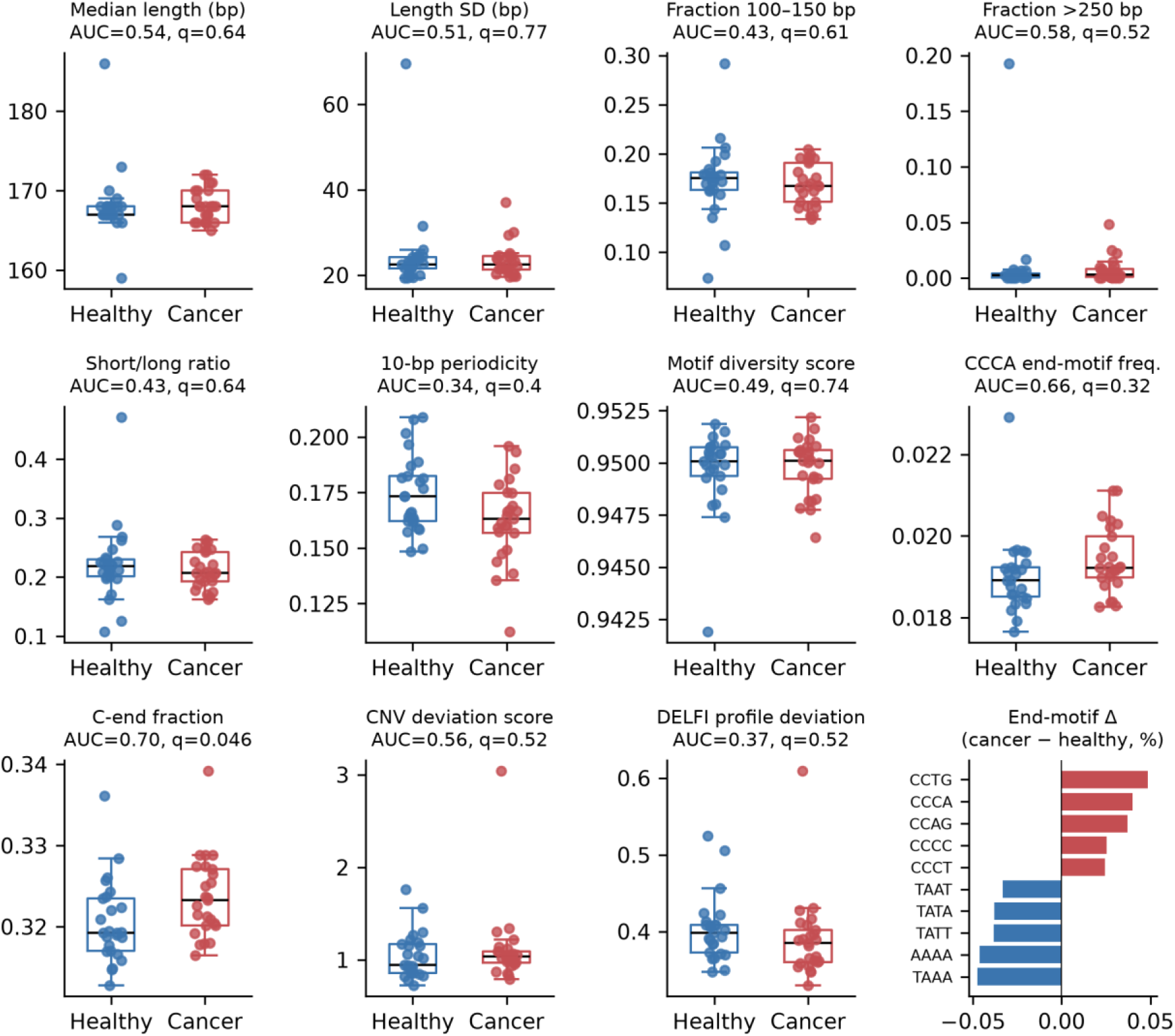
Genome-wide fragmentomic features without fragment-level GC control (breast). Boxplots with individual samples; titles give univariate AUC and BH-adjusted q values.

**Figure S4.**
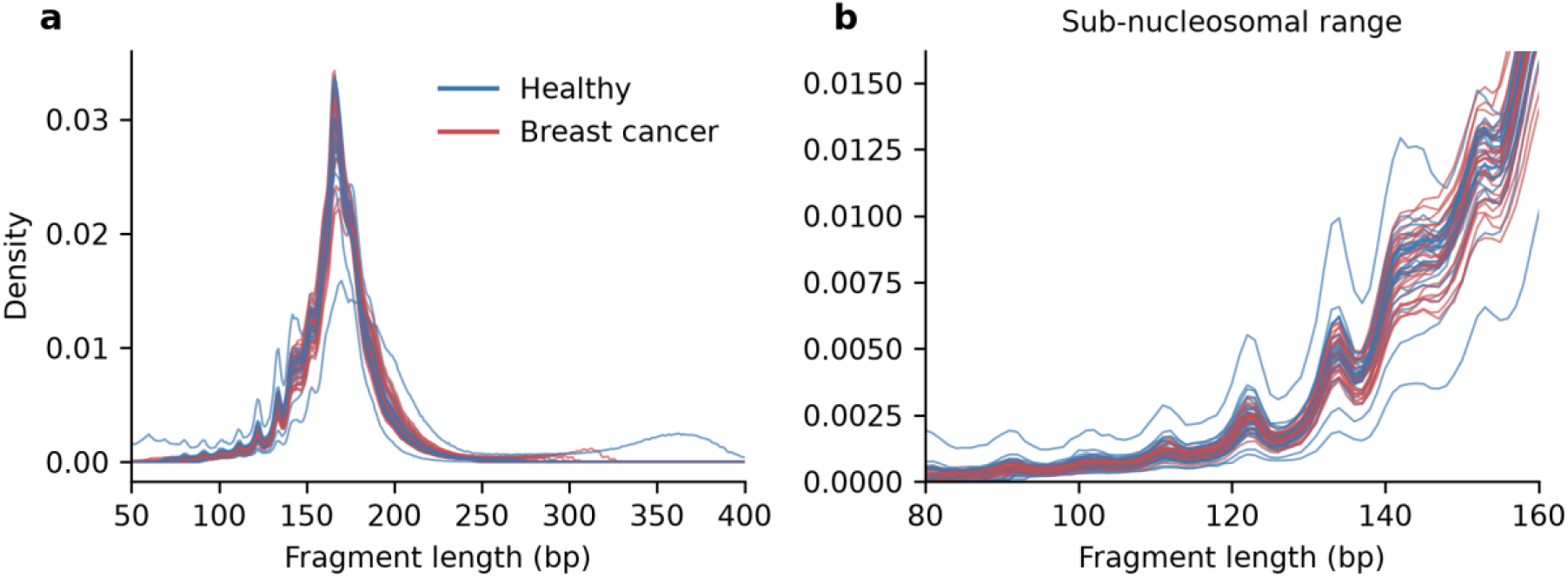
Fragment-length distributions in the breast cohort. (a) Length density per sample (blue, controls; red, breast cancer). (b) Sub-nucleosomal range.

## Supplementary Note 1 Liver cohort and per-pair values

### Cohort

There were 8 HCC cases (stage 1a, n = 2; 1b, n = 3; 2, n = 3; 5 male, 3 female; age 25–78 years) and 8 controls matched 1:1 for age, sex, smoking and alcohol use, from the same laboratory workflow as the breast cohort (13.3–50.0 M fragments per sample).

### Protocol

The protocol fixed before the liver data were unpacked specified three breast-trained, locked models: M1, the 11 fragmentomic features; M2, the multi-mapping fraction; M3, both. M1 gave AUC 0.77 (8/8 pairs) without and 0.44 (4/8) with fragment-level GC weighting; M2 gave 0.73 (5/8) and 0.69 (5/8); M3 gave 0.77 (6/8) and 0.48 (4/8). The heterochromatic-share model was added afterwards.

### Per-pair values (case / matched control)

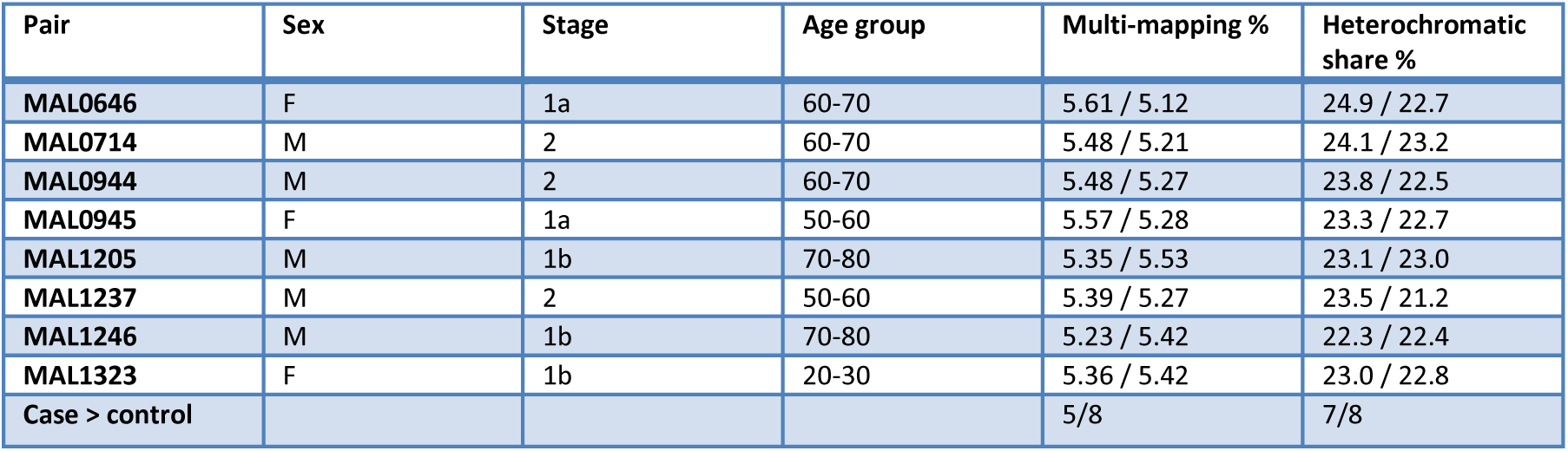

| Pair | Sex | Stage | Age group | Multi-mapping % | Heterochromatic share % |
|---|---|---|---|---|---|
| **MAL0646** | F | 1a | 60-70 | 5.61 / 5.12 | 24.9 / 22.7 |
| **MAL0714** | M | 2 | 60-70 | 5.48 / 5.21 | 24.1 / 23.2 |
| **MAL0944** | M | 2 | 60-70 | 5.48 / 5.27 | 23.8 / 22.5 |
| **MAL0945** | F | 1a | 50-60 | 5.57 / 5.28 | 23.3 / 22.7 |
| **MAL1205** | M | 1b | 70-80 | 5.35 / 5.53 | 23.1 / 23.0 |
| **MAL1237** | M | 2 | 50-60 | 5.39 / 5.27 | 23.5 / 21.2 |
| **MAL1246** | M | 1b | 70-80 | 5.23 / 5.42 | 22.3 / 22.4 |
| **MAL1323** | F | 1b | 20-30 | 5.36 / 5.42 | 23.0 / 22.8 |
| **Case > control** |  |  |  | 5/8 | 7/8 |

## Supplementary Note 2 GC-handling analyses of conventional features

The four analyses in Table 4 were run in the order listed. Analysis (ii) was added after analysis (i) had been completed in both cohorts, and analyses (iii) and (iv) followed a written protocol fixed before they were computed, including an addendum for the 10 M run. All four are reported.

Per-sample values for analyses (i) and (ii) are in Tables S1 and S2, in separate uncorrected and GC-weighted columns; within-fold residualisation results are in Table S3; relative-feature results are summarised in Table 4.

## Notes

### Competing Interest Statement

The authors have declared no competing interest.

### Author Declarations

This study used only openly available data: CRAG plasma cfDNA fragment files (Zenodo record 6914806, https://zenodo.org/records/6914806), TCGA BRCA whole-genome bisulfite data reprocessed by Zhou et al. (https://zwdzwd.github.io/pmd), Roadmap Epigenomics E066 methylation data (https://egg2.wustl.edu/roadmap/data/byDataType/dnamethylation/) and UCSC hg19 annotations (https://hgdownload.soe.ucsc.edu/goldenPath/hg19/).

### Summary of Updates

Misspelled the author names and organization name, so corrected that information.

